# Prognostic implications of splenomegaly in BCMA-directed CAR T-Cell therapy for relapsed myeloma

**DOI:** 10.1101/2025.01.31.25321490

**Authors:** Thomas Wiemers, Maximilian Ferle, Jonas Ader, Veronika Sotikova, David Fandrei, Nora Grieb, Luise Fischer, Patrick Born, Heike Weidner, Song Yau Wang, Madlen Jentzsch, Georg-Nikolaus Franke, Carmen Herling, Klaus Metzeler, Marco Herling, Simone Heyn, Timm Denecke, Kristin Reiche, Uwe Platzbecker, Vladan Vucinic, Hans-Jonas Meyer, Thomas Neumuth, Maximilian Merz

## Abstract

B-cell maturation antigen (BCMA)-directed chimeric antigen receptor (CAR) T-cell therapy has shown promise for patients with relapsed or refractory multiple myeloma (RRMM), yet its use is often complicated by adverse events such as cytokine release syndrome and hematologic toxicities. This study evaluates the prognostic significance of spleen size, measured via computed tomography (CT), in predicting clinical outcomes for RRMM patients undergoing CAR T-cell therapy. We retrospectively analyzed data from patients treated with Idecabtagene vicleucel or Ciltacabtagen autoleucel (N=73) and show that enlarged spleen size is associated with severe and prolonged thrombocytopenia, higher metabolic tumor volumes, and elevated soluble BCMA (sBCMA) levels. Our findings indicate that splenomegaly (>340 cm³) is an independent prognostic marker for both progression-free and overall survival, outperforming markers such as sBCMA levels, EASIX, and CAR- Hematotox scores. These results suggest that spleen size could enhance risk stratification in CAR T-cell therapy for RRMM patients, offering a readily accessible tool for guiding monitoring strategies and healthcare resource management.

## Introduction

B-cell maturation antigen (BCMA)-directed CAR T-cell therapy has emerged as a promising therapeutic option for patients with relapsed or refractory multiple myeloma (RRMM), offering deep and durable responses in a substantial proportion of patients. However, despite its efficacy, CAR T-cell therapy is associated with various adverse events, including cytokine release syndrome (CRS), neurotoxicity, and hematologic toxicities that pose a significant challenge post-CAR-T therapy, increasing the risk of infections, bleeding, and treatment delays (1–6).

Several clinical scoring systems, such as CAR-HEMATOTOX and EASIX, have been developed to predict the occurrence of adverse events like cytopenias, CRS and survival based on patient- and disease-related factors (7,8). Despite its role as a key modulator of hematopoiesis and immune regulation, the impact of spleen size has not yet been investigated in the contact of CAR T-cell therapies for RRMM (9,10). Importantly, splenomegaly or altered splenic function may exacerbate cytopenias by sequestering platelets and leukocytes at increased rates or by driving inflammatory pathways that impair bone marrow recovery (11).

In this study, we present a comprehensive longitudinal analysis evaluating the impact of multiple parameters, including biomarkers, adverse effects and scoring systems on clinical outcomes in a cohort of patients with RRMM undergoing CAR T- cell therapy. Specifically, we propose computed tomography (CT)-measured 3D spleen size as a novel prognostic marker, by demonstrating strong association with early disease progression and thrombocytopenia and superior predictive performance compared to existing scoring systems in our cohort, highlighting its potential utility for risk stratification and therapeutic success.

## Methods and Materials

### Patient selection and CAR T-cell therapy

The research presented in this study was conducted in line with the Declaration of Helsinki and was approved by the ethics committee of the University Hospital Leipzig (361/22-ek). Patients with RRMM undergoing treatment at the Department of Hematology, Hemostaseology and Cellular Therapy at University Hospital Leipzig were recruited after giving written informed consent and were not compensated for their participation. All data was analyzed retrospectively. Prior to CAR T-cell administration all patients underwent lymphodepleting therapy (LDC) with cyclophosphamide and fludarabine. Fludarabine dose was reduced to 50% in case of reduced glomerular filtration rate (GFR) <60 mL/min/1.73m^2^ and patients with severe renal impairment (<30 mL/min/1.73m^2^) received bendamustine. Three to five days after LDC, CAR T-cells were administered intravenously according to manufacturer instructions. All patients received bridging therapy (BT) with either bispecific antibodies, anti-CD38-based, anti-SLAMF7-based or conventional chemotherapy during CAR T-cell therapy.

### Imaging and Spleen Morphometry

Imaging was performed in a clinical setting using CT scanners (Ingenuity 128, iCT 256, Philips, Hamburg, Germany). The last CT exam before CAR T-cell therapy and the first staging CT after the therapy were analyzed. Intravenous administration of an iodine-based contrast agent (90 mL Imeron 400 MCT, Bracco Imaging Germany GmbH, Konstanz, Germany) was applied at a rate of 4.0 mL/s via a peripheral venous line. The software package from Philips (Intellispace portal, version 11; Philips, Philips Health System, Hamburg, Germany) was used to calculate the spleen volume (referred to as *spleen size*). The contour of the region of interest was located along the contour of the spleen semi-automatically. One trained radiology resident with 5 years of experience in radiology performed all measurements blinded to the clinical outcome.

### Risk Scoring

The CAR-HEMATOTOX Score was assessed as in previous studies using pre-LDC values of Platelet Count (PC), Absolute neutrophil count (ANC), Hemoglobin (Hb), Ferritin, and CRP. A score ≥2 was considered as high (12). Simultaneously, the EASIX Score was derived by calculating lactate dehydrogenase (LDH, U/L) × creatinine (mg/dL) / PC (G/L) prior to LDC and analyzed after applying log2 transformation (13,14). Clinical Scores were calculated by a trained physician blinded to the clinical outcome.

ANC and PC were monitored in all patients and scored according to the Common Terminology Criteria for Adverse Events (CTCAE v5). Thrombocytopenia was defined as a non-transient PC <150 G/L, confirmed by at least two measurements. Severe thrombocytopenia was classified as any grade 3 or 4 event, indicated by a PC <50 G/L and PC <25 G/L, respectively. Neutropenia was defined as a non- transient ANC <1.5 G/L, measured at least twice in succession. The severity of neutropenia was determined using the short and long immune effector cell- associated hematotoxicity (ICAHT) grading (15). Short ICAHT grading was based on the duration of severe neutropenia (ANC <0.5 G/L or <0.1 G/L) within the first 30 days post-treatment, while late ICAHT grading considered the severity of neutropenia (ANC<1.5; <1.0; <0.5 or <0.1 G/L) beyond 30 days post-CAR T-cell administration. Prolonged cytopenias were defined as neutropenia or thrombocytopenia persisting longer than 30 days post-treatment (16).

CRS and ICANS were graded based on the consensus criteria established by the American Society for Transplantation and Cellular Therapy (ASTCT) (17). The occurrence of infections was evaluated according to the CTCAE v5 criteria. Responders were defined as patients achieving PR or better according to IMWG criteria by month 6 after CAR T-cell infusion (18). Kaplan-Meier survival analyses were conducted from the time of CAR T-cell infusion to death from any cause (overall survival, OS) or to the progression of multiple myeloma (progression-free survival, PFS).

### Statistics

All results of this study were generated using the Python (version 3.10) programming language. We computed Spearman’s correlation coefficient to assess the relationship between variables using the scikit-learn package (version 1.5.2) package. Receiver operating characteristic analyses and computation of Area under ROC (AUROC) values done using scikit-learn as well. Significance testing of two populations was done using Mann-Whitney-U and Wilcoxon signed-rank tests where appropriate (refer to Figure legends) using the SciPy package (version 1.14.1). Survival analysis, Hazard analysis, and logrank significance tests were done using the lifelines package (version 0.29.0). Confidence intervals around Kaplan-Meier- Curves were estimated by consideration of right-censoring using Greenwood’s Exponential formula. P-values of <0.05 and <0.001 were considered statistically significant, as indicated in the figure legends respectively. Splenomegaly cut-off was calculated by optimizing the logrank statistic between two groups, which were stratified through thresholding. Here, only the data between the 15^th^ and 85^th^ percentile were considered to prevent edge artifacts. For both PFS and OS this resulted in an optimal threshold value of 340 cm³. Notably, this cut-off is of close to equal magnitude of those estimated by other groups using entirely different methodology (19). Cut-offs for other markers used in survival analysis were computed in the same manner.

## Results

### Clinical characteristics and outcomes following anti-BCMA CAR T-cell therapy

In this retrospective analysis, included 73 patients with RRMM, who were treated with the commercial CAR T-cell products Idecabtagene vicleucel (Ide-cel, N=35) or Ciltacabtagene autoleucel (Cilta-cel, N=38) after three lines of therapy or more between 2021 and 2024 at the Department of Hematology, Hemostasis, Cellular Therapy and Infectiology at the University Hospital of Leipzig. A comprehensive description of the patient cohort is presented in Table 1. CT scans performed before CAR T-cell administration were available for 64 (87.7%), while post-treatment scans for 58 (79.5%) patients (Figure 1A). These scans were used to assess the impact of spleen size in the context of CAR T-cell therapy. Median patient age was 64 (range: 31-78), with 31 (42.4%) women and 42 (57.6%) men. R-ISS stage III was detected in 15 (20.5%) Patients, 40 (54.3%) exhibited high-risk cytogenetic aberrations. Extramedullary disease affected 19 (26.0%), including bone-associated masses in 6 (6.82%) cases and extraosseous soft tissue masses in 13 (17.8%) patients. The median follow-up for PFS and OS was 8.2 and 6.1 months, respectively. The distribution of treatments that preceded CAR T-cell treatment is presented in Figure 1B. Triple-class refractory disease refractory disease was observed in 50 (71.4%) and penta-drug in 13 (18.6%) of cases. The median of therapies prior to CAR T-cell therapy was 7 (1-17). The overall response rate (ORR) to CAR T-cell therapy was 82.2% and complete response (CR) was achieved by 45 (61.6%) patients. Importantly, 3 (4.1%) patients of our cohort received CAR T-cell therapy twice.

**Figure 1.**
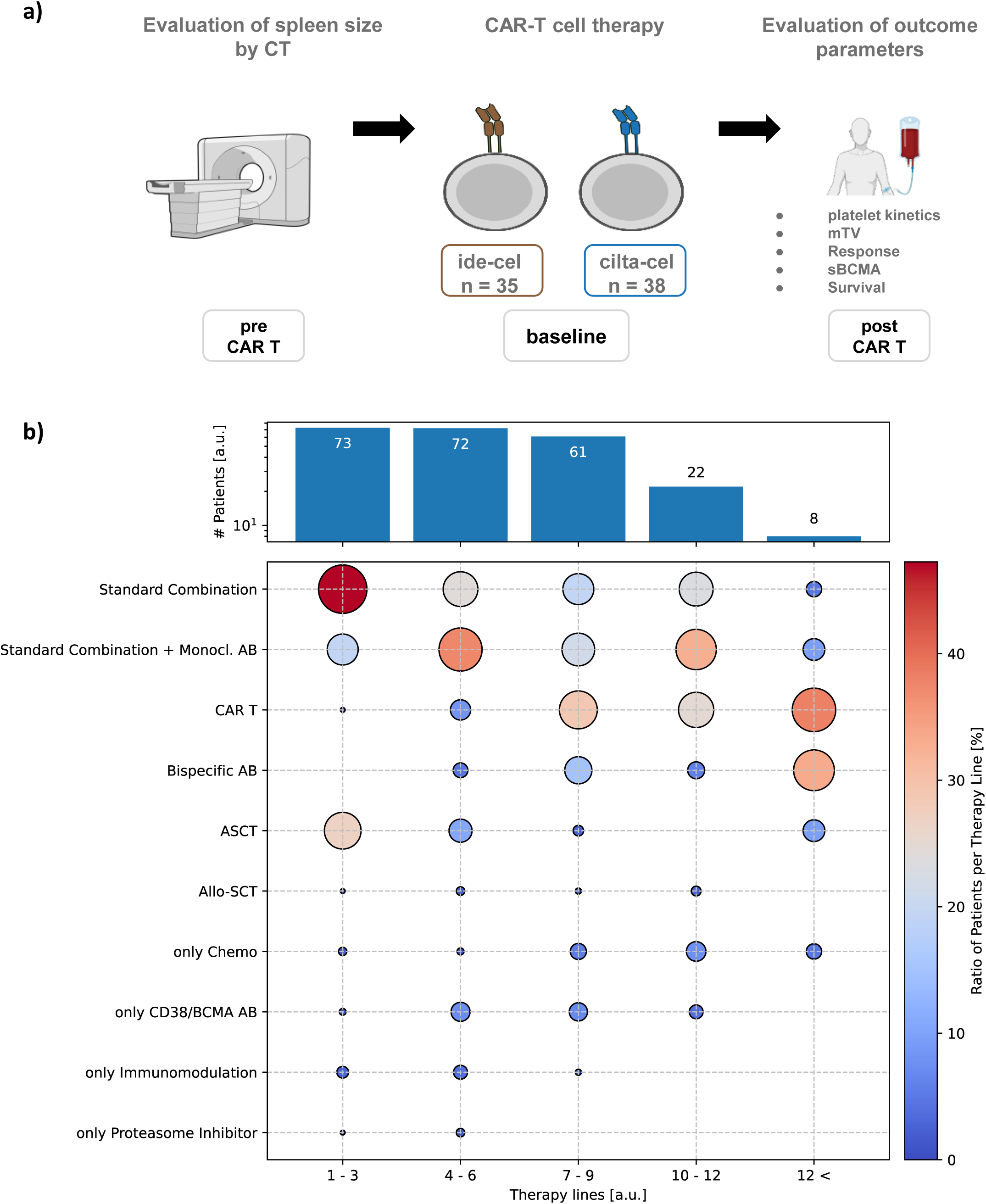
Evaluation of Splenomegaly prior to CAR T-Cell Therapy and Summary of Cohort Treatment History. (**a**) Workflow of the study. Spleen size is assessed using computed tomography (CT) in patients before receiving CAR T-cell therapy. Two different CAR T products are administered ide-cel (n = 35 patients) and cilta-cel (n = 38 patients). Subsequently, various outcome parameters were evaluated. (**b**) Summary of the cohort’s treatment history prior to CAR T-cell administration. The upper bar graph displays the total number of patients receiving different treatment regimens within the respective therapy lines. The colored circles in the lower section represent the ratio of patients per therapy line prior to CAR T treatment, with color intensity indicating the proportion (% ratio of patients; ranging from 0% to 40%) across various prior therapies such as Standard Combination, Monoclonal Antibody (Monocl. AB), CAR T, Bispecific Antibody (AB), Autologous Stem Cell Transplantation (ASCT), and others. Multiple therapy lines were condensed (1-3, 4-6, 7-9, 10-12, and >12) for conciseness.

**Table 1:** Patient characteristics.

|  | **total (n=73)** |
| --- | --- |
| **Male - no. (%)** | 42 (57,6%) |
| **Female - no. (%)** | 31 (42,4%) |
| **Age - median (range)** | 64 (31-78%) |
| **R2-ISS III - no. (%)** | 15 (20,5%) |
| **High-risk cytogenetics (del 17, +1q, t4;14, t(14;16)) - no. (%)** | 40 (54,3%) |
| **Extramedullary disease - no. (%)** | 19 (26,0%)* |
| Bone associated masses - no. (%) | 6 (6,82%)* |
| extraosseus soft tissue masses - no. (%) | 13 (17,8%)* |
| **CAR after CAR treatment** | 3 (4,1%) |
| **Prior therapy lines - median (range)** | 7 (1-17) |
| Lenalidomide exposed - no. (%) | 69 (98,6%)* |
| Pomalidomide exposed - no. (%) | 57 (81,4%)* |
| Bortezomib exposed - no. (%) | 69 (98,6%)* |
| Carfilzomib exposed - no. (%) | 54 (77,1%)* |
| Daratumumab exposed - no. (%) | 70 (100%)* |
| triple class refractory - no. (%) | 50 (71,4%)* |
| penta class refractory - no. (%) | 13 (18,6%)* |
| Response Rate - no. (%) | 60 (82,2%) |
| Complete Response archived - no. (%) | 45 (61,6%) |
| **Products/ Infusion** |  |
| ice-cel | 35 (47,9%) |
| cilta-cel | 38 (52,1%) |

\*CAR after CAR not counted,n=70

**Table 2:** Adverse effects and prognostic evaluations.

|  |  |
| --- | --- |
| - | **total (n=73)** |
| **CRS any grade - no. (%)** | 54 (73,9%) |
| I° | 39 (53,4%) |
| II° | 14 (19,2%) |
| III° | 1 (1,3%) |
| **ICANS any grad -no. (%)** | 6 (8,2%) |
| I° | 5 (6,8%) |
| II° | 1 (1,3%) |
| **Medication - no. (%)** | - |
| Tocilizumab received | 21 (28,7%) |
| **Infections of any grade - no. (%)** | 37 (50,7%) |
| 0° | 36 (49,3%) |
| I° | 2 (2,7%) |
| II° | 8 (10,9%) |
| III° | 19 (26,0%) |
| IV° | 4 (5,8%) |
| V° | 4 (5,5%) |
| **Cause of infection** | - |
| viral | 6 (8,2%) |
| bacterial/ catheter related infection | 28 (38,4%) |
| fungal | 1 (1,4%) |
| unknown | 12 (16,4%) |
| **Thrombopenia any grade - no. (%)** | 60 (82,2%) |
| Thrombopenia ≥ CTC 3 - no. (%) | 19 (26,0%) |
| median duration d (range) | 31 (3-569) |
| Thrombopenia ≥30 d - no. (%) | 25 (34,2%) |
| Preexisting Thrombopenia - no. (%) | 39 (53,4%) |
| **Neutropenia any grade - no. (%)** | 73 (100%) |
| median duration d (range) | 14 (1-119) |
| e -ICAHT ≥ 3 - no. (%) | 7 (9,6%) |
| I - ICAHT ≥ 3 - no. (%) | 5 (6,8%) |
| Neutropenia ≥ 30 days - no. (%) | 11 (15,1%) |
| cumulative days of neutropenia <500/ul (range) in 30 days after CAR T - no. (%) | 6 (0-30) |
| Preexisting Neutropenia - no. (%) | 39 (53,4%) |
| **neutrophil recovery type** | - |
| quick - no. (%) | 45 (61,6%) |
| intermittent - no. (%) | 21 (28,8%) |
| aplastic - no. (%) | 4 (5,4%) |
| not evaluable - no. (%) | 3 (4,1%) |
| **Predictive scores and values** | - |
| EASIX ≥ 1,87 - no. (%) | 12 (16,4%) |
| CAR Hematotox ≥ 2 - no. (%) | 30 (41,1%) |
| Spleen size > 340 cm³ pre CAR T - no (%) | 15 (20,5%) |
| sBCMA > 175 ng/mL at leukapheresis - no (%) | 19 (26,0%) |
| **Secondary malignancies** | - |
| Myelodysplastic syndrome (MDS) | 1 (1,4%) |

CRS of any grade occurred in 54 patients (73.9%), with the majority experiencing grade I CRS (53.4%), followed by grade II (19.2%) and grade III (1.3%). Immune effector cell-associated neurotoxicity syndrome (ICANS) was less frequent, occurring in 6 patients (8.2%), with grade I in 5 patients (6.8%) and grade II in 1 patient (1.3%). In terms of medication for CRS, 21 patients (28.7%) received tocilizumab. Infections were reported in 37 patients (50.7%), while infections of grades I, II, III, IV, and V were observed in 2.7%, 10.9%, 26.0%, 5.8%, and 5.5% of the cohort, respectively. The causes of infection were primarily bacterial or catheter- related (38.4%), followed by viral (8.2%), fungal (1.4%), and unknown origins (16.4%).

### Hematologic toxicities after CAR T-cell infusion

Thrombocytopenia of any grade was observed in 60 patients (82.2%), with grade ≥3 thrombocytopenia in 19 patients (26.0%) and a median duration of 31 days (range: 3–569). Prolonged thrombocytopenia (≥30 days) was seen in 25 patients (34.2%). Neutropenia was universal in this cohort, with a median duration of 14 days (range: 1–119). Prolonged neutropenia (≥30 days) was observed in 11 patients (15.1%). In order to combine severity and duration of neutropenia ICAHT grading was utilized. Early ICAHT (e-ICAHT) grade >2 occurred in 7 patients (9.6%), while late ICAHT (l- ICAHT) grade >2 was seen in 5 patients (6.8%). Patterns of neutrophil recovery varied, with 45 patients (61.6%) experiencing quick recovery, 21 patients (28.8%) having intermittent recovery, and 4 patients (5.4%) showing an aplastic recovery pattern.

### Spleen size is an independent prognostic parameter for PFS and OS

To elucidate the impact of clinical and biological markers on the outcomes of anti- BCMA-directed CAR T-cell therapy in RRMM, we focused on identifying key predictors of hematologic toxicities and overall treatment efficacy. Consequently, we explored the Spearman correlations (Figure 2A) of clinical markers included in our assessment to pinpoint clinically relevant associations that could inform risk stratification strategies.

**Figure 2.**
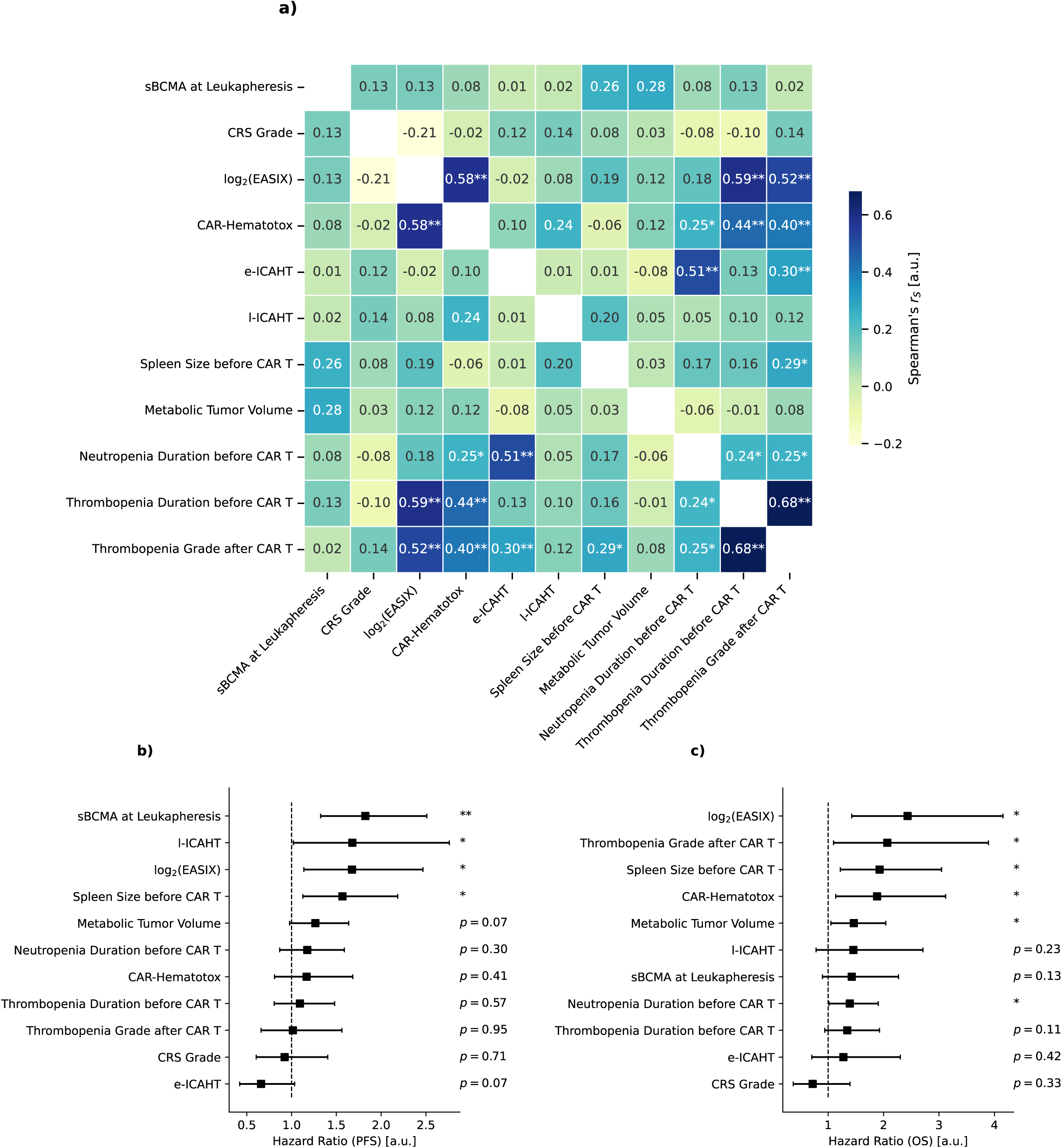
Analysis of Prognostic and Outcome Parameters in CAR T Therapy. (**a**) Correlation matrix depicting Spearman’s rank correlation coefficients (*rS*) among various prognostic and outcome parameters measured in patients prior to and during CAR T therapy. Each cell displays the correlation coefficient, with shading indicating the strength and direction of the correlation (values range from -0.2 to 0.6, as represented by the color scale). Significant correlations are indicated by asterisks (two asterisks: p < 0.001; one asterisk: p < 0.05). (**b**) Forest plot illustrating the hazard ratios (HR) with corresponding confidence intervals for factors influencing progression-free survival (PFS) post-CAR T therapy. Statistical significance is noted for specific variables, with asterisks marking relevant p-values (analogous to panel a). (**c**) Forest plot presenting HR for overall survival (OS) related to the same predictors.

The correlation analysis revealed several associations. High EASIX scores were strongly associated with severe thrombocytopenia (*rS* = 0.52, p < 0.001) and duration of thrombocytopenia at baseline (*rS* = 0.59, p < 0.001), demonstrating the reliability of EASIX as a marker for hematological toxicity. Similarly, the CAR-HEMATOTOX scores showed significant correlations with the duration of thrombocytopenia (*rS* = 0.44, p < 0.001), underscoring its relevance in predicting hematologic adverse effects. Interestingly, while spleen size demonstrated moderate correlation with thrombocytopenia grade after therapy (*rS* = 0.29, *rS* < 0.05), it remained independent of other commonly evaluated markers such as soluble BCMA (sBCMA) levels, EASIX, and CAR-HEMATOTOX scores (Figure 2A). Further statistical analysis reinforced the prognostic relevance of spleen size. While EASIX was confirmed to significantly influence both PFS (Figure 2B, HR = 1.67, 95% CI 1.14 - 2.47, p < 0.05) and OS (Figure 2C, HR = 2.43, 95% CI 1.43 - 4.16, p < 0.05), markers such as sBCMA (HR = 1.82, 95% CI 1.32 - 2.51, p < 0.001), and CAR-HEMATOTOX (HR = 1.88, 95% CI 1.14 - 3.12, p < 0.05) only influenced PFS or OS respectively. Spleen size notably demonstrated a high association with both PFS (HR = 1.57, 95% CI 1.13 - 2.19, p < 0.05) and OS (HR = 1.93, 95% CI 1.22 - 3.05 p < 0.05). Here, spleen size emerged as a strong predictor, with hazard ratios comparable to those of traditional markers like BCMA, EASIX, and CAR-HEMATOTOX.

The identification of spleen size as a critical and independent marker strongly influencing both PFS and OS in patients undergoing CAR T-cell therapy prompted us to analyte the clinical implications of increased spleen sizes in greater detail.

### Spleen size prior to CAR T-cell therapy is associated with thrombocytopenia, metabolic CR and tumor burden

To further delineate the predictive value of spleen size on outcomes after CAR T- cell therapy, we conducted a series of analyses exploring its relationship with post- treatment complications and response metrics.

Across the cohort, no significant changes in spleen size (Figure 3A-B) were observed before and after therapy (p=0.278, Figure 3C). Notably, subgroup analyses examining patients with higher-grade thrombocytopenia, elevated e- ICAHT, l-ICAHT, prolonged thrombocytopenia and neutropenia revealed no significant changes in spleen size before and after therapy (Supplementary Figure 1).

**Figure 3.**
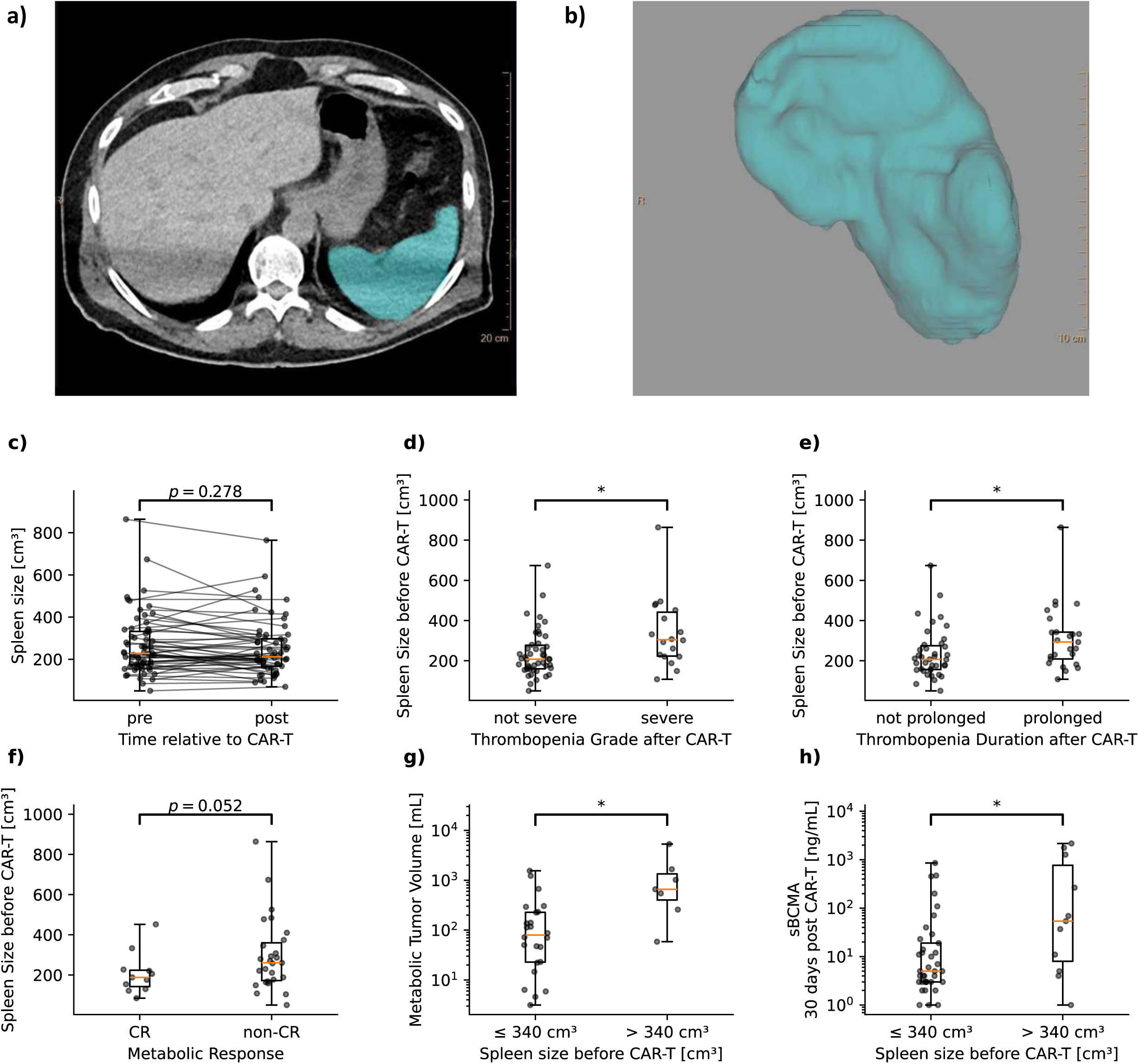
Relation of spleen size to CAR T-cell therapy outcomes. (**a**) Axial computed tomography (CT) image depicting the volume of the spleen prior to CAR T-cell therapy, with the highlighted region indicating the spleen. (**b**) Three- dimensional reconstruction of the spleen from scans shown in panel a, providing a visual representation of splenic morphology. (**c**) Spleen size before and after CAR T therapy, showing no significant change (p = 0.278) in volume measured over time. (**d**) Comparison of spleen size before CAR T therapy categorized by thrombocytopenia grade after therapy. Significant differences were observed, with spleen size being greater in patients classified as having severe thrombocytopenia (p < 0.05). (**e**) Comparison of spleen size before CAR T therapy categorized by the duration of thrombocytopenia after therapy. Results indicated that patients with prolonged thrombocytopenia had larger spleens compared to those without prolonged duration (p < 0.05). (**f**) Evaluation of spleen size before CAR T therapy in relation to metabolic response (complete response (CR) vs. non-CR), revealing a weakly significant trend (p = 0.052). (**g**) Relationship between spleen size before CAR T therapy and metabolic tumor volume (mTV) in patients lacking CR (mTV > 0 mL). Significant differences (p < 0.05) were found between patients with splenomegaly (spleen size > 340 cm³) and those with normal sized spleens. (**h**) sBCMA activity measured 30 days post CAR T therapy in relation to the spleen size prior to treatment, demonstrating significant differences (p < 0.05) between patients with splenomegaly and those without. Statistical significance was assessed using one-sided Mann-Whitney-U tests.

However, patients who experienced severe (Figure 3D) as well as prolonged (Figure 3E) thrombocytopenia after treatment had significantly larger spleens prior to therapy compared to those without or non-severe thrombocytopenia (p < 0.05, respectively). Also, Patients who did not achieve a metabolic complete response (metCR) in PET/CT post-treatment had larger spleens compared to those who achieved a metCR (p = 0.052, Figure 3F). Moreover, among patients lacking a metabolic CR, those with splenomegaly (spleen size > 340 cm³; refer to methods section *Statistics* for rationale) demonstrated significantly greater metabolic tumor volumes than those with normal-sized spleens (p < 0.05, Figure 3G). Furthermore, our analysis indicated that patients with splenomegaly had significantly higher sBCMA levels 30 days after receiving CAR T-cell therapy compared to those with normal spleen sizes (p < 0.05, Figure 3H).

The significant associations between enlarged spleens and both severe and prolonged thrombocytopenia, higher metabolic tumor volumes, and elevated sBCMA levels underscore the potential of pre-treatment spleen size as a critical marker for outcomes following CAR T-cell therapy.

### Splenomegaly prior to CAR T-Cell infusion predicts treatment response and survival, outperforming EASIX and CAR-HEMATOTOX

In our analysis of predictive markers for CAR T-cell therapy survival outcomes, illustrated in Figure 4, spleen size emerged as a paramount indicator.

**Figure 4.**
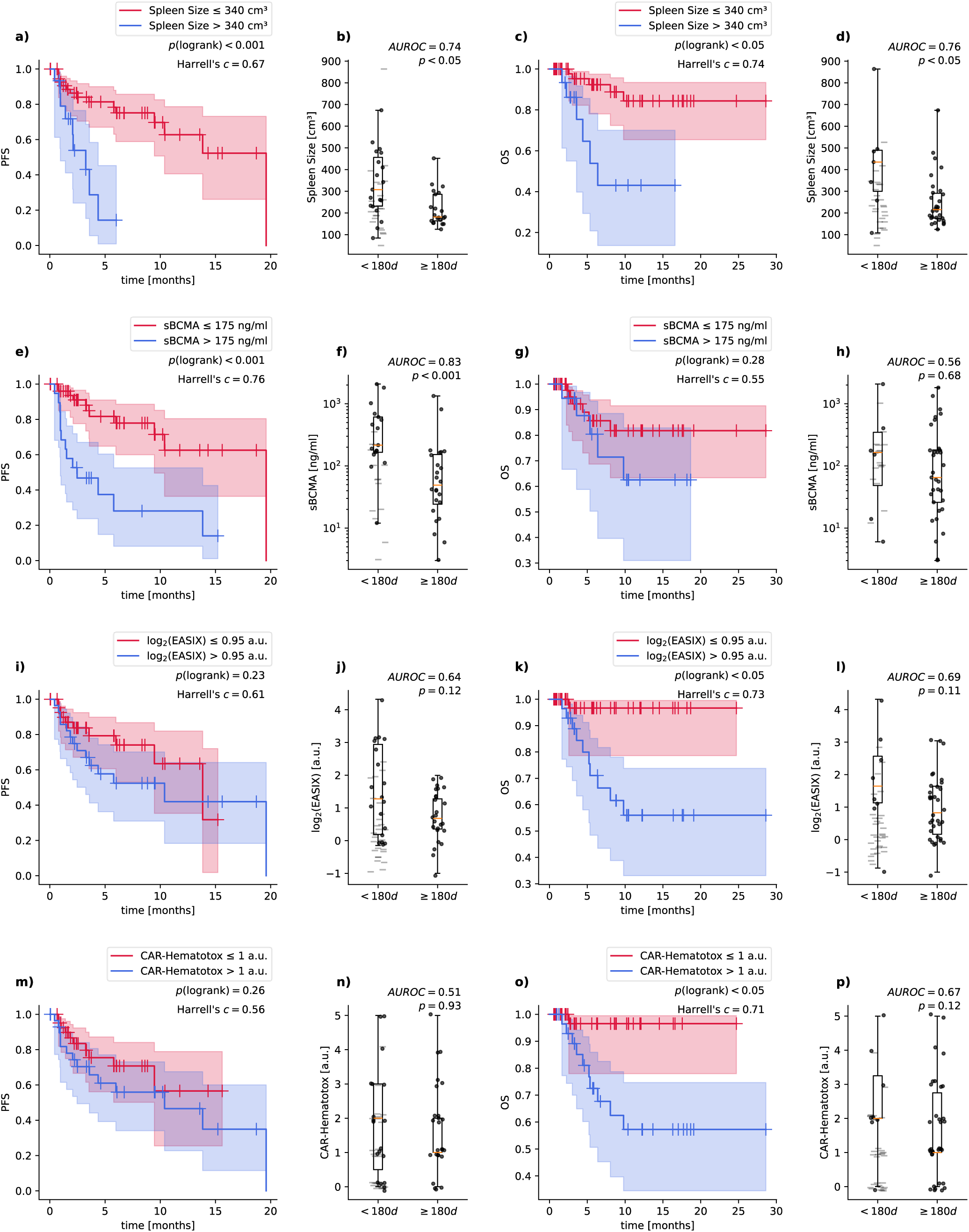
Survival analysis and assessment of spleen size compared to various prognostic markers. (**a-d**) Evaluation of the influence of spleen size on progression-free survival (PFS, **a**) and overall survival (OS, **c**). Kaplan-Meier curves are displayed for patients stratified by spleen size ≤ 340 cm² (red) and > 340 cm² (blue). The area under the receiver operating characteristic curve (AUROC) was evaluated for survival of < 180 days and ≥ 180 days with respect to PFS (**b**) and OS (**d**). Dots indicate events, while transparent dashes indicate censoring. (**e-h**) Assessment of prognostic significance of sBCMA levels. Kaplan-Meier curves illustrate PFS (**e**) and OS (**g**) stratified by sBCMA ≤ 175 ng/ml (red) versus > 175 ng/ml (blue), while AUROC analyses were performed in analogy to panels b and d for PFS (**e**) and OS (**h**). (**i-l**) Impact of log2(EASIX) levels on PFS (**i**) and OS (**k**). Patients are stratified into log2(EASIX) ≤ 0.95 (red) and > 0.95 (blue). AUROC analyses were performed in analogy to panels b and d for PFS (**j**) and OS (**l**). (**m-p**) Exploration of the association between CAR- Hematotox scores and PFS (**m**) and OS (**o**). Kaplan-Meier curves depict survival profiles based on CAR-Hematotox ≤ 1 (red) versus > 1 (blue). AUROC analyses were performed in analogy to panels b and d for PFS (**n**) and OS (**p**), respectively. All survival analyses include corresponding p-values, Harrell’s c-index values, and AUROC scores where applicable.

For PFS, patients with Splenomegaly (spleen size > 340 cm³; See methods section *Statistics* for rationale) exhibited notably worse outcomes compared to those with smaller spleens, as evidenced by Kaplan-Meier survival analyses (p(logrank)<0.001, c=0.67, Figure 4A) and supported by a high area under the receiver operating characteristic curve (AUROC=0.74, p<0.05, Figure 4B) for predicting survival times of less than 180 days. In comparison, sBCMA levels showed a slightly superior predictive power for survival (p(logrank)<0.001, c=0.76, Figure 4E) and discriminative performance for outcomes within 180 days (AUROC=0.83, p<0.001, Figure 4F). However, established markers such as EASIX and CAR-HEMATOTOX scores were less effective, with EASIX showing only moderate predictive value (Figure 4I-J) and CAR-HEMATOTOX demonstrating low performance (Figure 4M-N) in the same context.

For OS, spleen size not only maintained its predictive relevance but outperformed all other studied markers. Splenomegaly demonstrated even higher utility for risk stratification in the context of OS (p(logrank)<0.05, c=0.74, Figure 4C) and predicting survival events within 180 days (AUROC=0.76, p<0.05, Figure 4D). This was notably higher than the predictive values obtained for sBCMA, EASIX, and CAR-HEMATOTOX. Interestingly, while sBCMA levels did not demonstrate significant utility in the context of OS (Figure 4G-H), both EASIX and CAR-HEMATOTOX scores showed only modest AUROC values for late outcomes (AUROC=0.69 and 0.67 respectively, Figures 4L and 4P).

These results collectively highlight the promising role of spleen size, particularly when exceeding 340 cm³, in risk stratification for both PFS and OS. The demonstrated results strongly imply that assessment of spleen size would increase accuracy of prognostic models used in CAR T-cell therapy for multiple myeloma.

## Discussion

In this study, we demonstrated the implications of splenomegaly for risk stratification of patients with RRMM undergoing CAR T-cell therapy. The assessment of spleen characteristics in imaging modalities such as CT and MRI has been explored as a prognostic marker across various diseases, including hematological malignancies and solid tumors (20–22). In hematologic conditions, splenic enlargement often correlates with disease burden or immune activation as highlighted in the context of myelofibrosis and chronic lymphocytic leukemia (CLL) (20,21,23). In patients with multiple myeloma, loss of spleen visualization (LOV) was associated with poor prognostic outcomes, including reduced OS and PFS in whole-body diffusion-weighted imaging (WB-DWI) and ^68^Ga-Pentixafor-PET/CT (24–26). However, to the best of our knowledge, no study explored spleen size enlargement as a prognostic marker in the context of CAR T-cell therapy before. We demonstrated the utility of spleen size as a novel, independent prognostic parameter that is associated with higher risks of severe and prolonged thrombocytopenia, as well as lower metabolic response rates and elevated sBCMA and mTV levels. Compared to other established scores commonly used to assess the likelihood of hematologic toxicities, spleen size emerged as the most reliable marker for prognosis of survival outcomes in our cohort. Notably, spleen size is an easily accessible and, based on our evaluations, independent marker that can be evaluated prior to treatment.

It is well known that the majority of patients exhibit cytopenia after administration of CAR T-cells (27). Here, pre-existing splenomegaly may contribute to the duration and severity of thrombocytopenia as the spleen’s capacity to sequester platelets and other hematopoietic elements may exacerbate cytopenias following CAR T-cell therapy. In this context we could observe an association between spleen size and the severity and duration of thrombocytopenia after therapy. This phenomenon has been documented in other hematologic conditions, such as myelofibrosis, where splenic enlargement was associated with altered platelet dynamics and hematopoiesis after allogeneic stem cell transplantation (23). Severe and prolonged hematologic toxicities have already been identified as risk factors for lower PFS and OS in patients suffering from MM and other hematologic malignancies treated with CAR T-cells (4,5). Many studies observed reduced hematopoietic activity in the bone marrow of patients with cytopenia following CAR T-cell therapy (28,29). For example, a study examining a small cohort found that cytopenia was associated with hypoplasia in the bone marrow, indicating impaired hematopoiesis, which may contribute to the prolonged cytopenic state (30). Beyond the sequestration of platelets, the prognostic significance of the spleen size may be also linked to its role in immune cell trafficking and extramedullary hematopoiesis (EMH) (26,31,32). EMH can generally affect every organ system in the body as it has already been reported to be localized pulmonary, intracranial or paravertebral (33–35). Moreover, the ability of the spleen to switch into a hematopoietic organ after e.g. myeloablative chemotherapy has been demonstrated in-vitro (31,36). Thus, we hypothesize that spleen size may indirectly reflect EMH activity and reduced immune cell trafficking which may explain the reduction of PFS and OS observed in our cohort.

Several studies have highlighted the adverse impact of prolonged cytopenia on patient outcomes, including an increased risk of infections, progression, and poor survival rates (37). For example, prolonged cytopenia has been associated with both a higher incidence of CRS and a greater vulnerability to infections, particularly bacterial infections within the first 30 days of therapy. These complications in turn, contribute to worse OS and PFS (1,4). Based on our results, assessing spleen size could serve as a useful tool to inform the decision of administering CAR T-cell treatment and, along other established markers, may serve to identify high-risk patients, who would benefit from closer monitoring during treatment to facilitate early intervention in case of disease progression.

Moreover, in our cohort spleen size was the most powerful marker overall for estimating both PFS and OS, outperforming established markers like sBCMA, EASIX and CAR-HEMATOTOX. However, the observed differences in prognostic outcomes from sBCMA, EASIX and CAR-HEMATOTOX may also be attributed to different bridging regimens or the usage of different CAR T-cell products. It is important to note that, in recent studies focusing on EASIX and CAR-HEMATOTOX, ide-cel was generally used, with only a few patients in these cohorts treated with cilta-cel (7,8). In our study, half of the patients received cilta-cel, which has been associated with distinct toxicity and efficacy profiles compared to ide-cel, particularly regarding neurotoxicity and CAR-T cell expansion (38). This may promise enhanced generalizability as compared to other studies focusing only on ide-cel.

Taken together, our study provides strong evidence for the prognostic significance of spleen size as a novel, accessible, independent marker in patients with RRMM undergoing CAR T-cell therapy. By addressing the multifactorial nature of post-CAR T complications and their nuanced impact on survival outcomes, our findings improve the assessment of risk factors in an easily accessible setting. They also underscore the importance of developing individualized management strategies to optimize patient care and improve treatment outcomes. We could show that splenomegaly as a prognostic marker outperforms established biomarkers like sBCMA, EASIX, and the CAR-HEMATOTOX score, offering potential utility as a tool to refine patient monitoring strategies, leading to improved healthcare resource management and patient care.

## Acknowledgements

This work was funded by grants from the International Myeloma Society (IMS Research Grant 2023) and the German Research Foundation (SPP µbone) and EU HORIZON (CERTAINTY). The CERTAINTY project is funded by the European Union (Grant Agreement 101136379). Views and opinions expressed are however those of the authors only and do not necessarily reflect those of the European Union. Neither the European Union nor the granting authority can be held responsible for them.

The authors acknowledge the financial support by the Federal Ministry of Education and Research of Germany and by Sächsisches Staatsministerium für Wissenschaft, Kultur und Tourismus in the programme Center of Excellence for AI-research “Center for Scalable Data Analytics and Artificial Intelligence Dresden/Leipzig”, project identification number: ScaDS.AI The graphical abstract was partly generated using NIH BIOART, licensed under a NIAID Visual & Medical Arts. (7.10.2024) license.

Authors express their gratitude towards patients and their families who participated in our research.

## Author contributions

TW and MF analyzed data, designed figures and took lead in writing the manuscript. JA analyzed data.

DF and NG contributed to conceptualizing the analyses.

LF, HW, SYW, MJ, GNF, CH, KM, MH, SH managed patient care and data collection.

HJM and VS performed CT imaging and spleen morphometry analyses. TD, KR, UP, VV, TN and MM supervised the project.

HJM and MM devised the main conceptual ideas.

All authors discussed the results have approved of the manuscript.

## Data availability

The data used in this study is available upon reasonable request to.

---

Supplementary Figure 1 Changes in spleen size relative to CAR-T therapy across various different patient subgroups (**a**) Patients with thrombocytopenia greater than Grade 2 show no significant difference in spleen size (p = 0.330). (**b**) Patients with e-ICAHT greater than Grade 3 exhibit a non-significant change in spleen size (p = 0.185). (**c**) The cohort with I- ICAHT greater than Grade 2 also demonstrated no significant alteration in spleen size (p = 0.185). (**d**) Patients experiencing thrombocytopenia for more than 30 days indicate a non-significant change in spleen size (p = 0.206). (**e**) Neutropenic patients beyond 30 days show no significant change in spleen size (p = 0.330). (**f**) Patients with CTCAE infections greater than Grade 2 reveal no significant difference in spleen size (p = 0.798). Statistical significance was assessed using Wilcoxon signed-rank tests.

