## Supplementary figures and images for "Prognostic implications of splenomegaly in BCMA-directed CAR T-Cell therapy for relapsed myeloma"

### Supplemental Figure 1

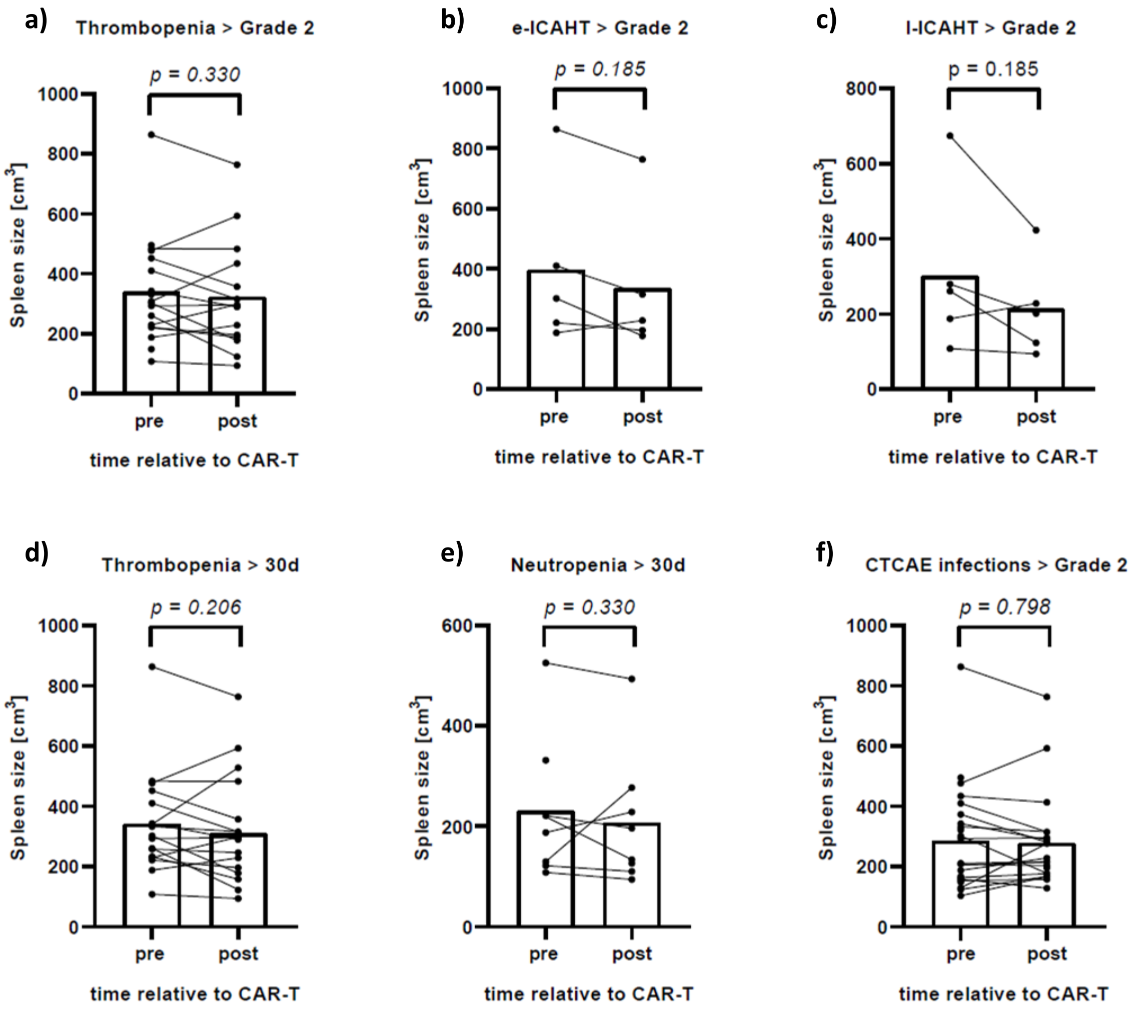
